## Supplementary material for "Getting to GRIPS with MR-Egger: modelling directional pleiotropy independently of allele coding": S1 Text

### Supporting information

#### 1. MR-GRIP estimator

The design matrix in MR-GRIP is

$$X=\left( \begin{matrix} 1 & \hat{\gamma}_{1}^{2} \\ 1 & \hat{\gamma}_{2}^{2} \\ \begin{matrix} \vdots\\ 1 \end{matrix} & \begin{matrix} \vdots\\ \hat{\gamma}_{m}^{2} \end{matrix} \end{matrix} \right)$$

(1)

with weight matrix

$$W=\left( \sum w_{j} \right)^{-1}diag\left( w_{1}, w_{2},\cdots,w_{m} \right)$$

(2)

where $w_{j}=\hat{\gamma}_{j}^{-2}\sigma_{Y_{j}}^{-2}$, and outcome vector

$$Y=\left( \begin{matrix} \hat{\Gamma}_{1}\hat{\gamma}_{1} \\ \hat{\Gamma}_{2}\hat{\gamma}_{2} \\ \begin{matrix} \vdots\\ \hat{\Gamma}_{m}\hat{\gamma}_{m} \end{matrix} \end{matrix} \right)$$

(3)

Standard theory gives the weighted least-squares estimates as $\left( \hat{\alpha}_{0},\hat{\beta} \right)^{T}=\left( X^{T}WX \right)^{-1}X^{T}WY$, with the causal effect explicitly

$$\hat{\beta}=\frac{\sum w_{j}\sum w_{j}\hat{\Gamma}_{j}\hat{\gamma}_{j}^{3}-\sum w_{j}\hat{\Gamma}_{j}\hat{\gamma}_{j}\sum w_{j}\hat{\gamma}_{j}^{2}}{\sum w_{j}\sum{w_{j}\hat{\gamma}}_{j}^{4}-\left( \sum w_{j}\hat{\gamma}_{j}^{2} \right)^{2}}$$

(4)

and the intercept

$$\hat{\alpha}_{0}=\frac{\sum w_{j}\hat{\gamma}_{j}^{4}\sum w_{j}\hat{\Gamma}_{j}\hat{\gamma}_{j}-\sum w_{j}\hat{\Gamma}_{j}\hat{\gamma}_{j}\sum w_{j}\hat{\gamma}_{j}^{2}}{\sum w_{j}\sum{w_{j}\hat{\gamma}}_{j}^{4}-\left( \sum w_{j}\hat{\gamma}_{j}^{2} \right)^{2}}$$

(5)

The standard errors are respectively

$$se\left( \hat{\beta} \right)=\left( \frac{\sum w_{j}\sum w_{j}\left( \hat{\Gamma}_{j}\hat{\gamma}_{j}-\hat{\alpha}_{0}-\hat{\beta}\hat{\gamma}_{j}^{2} \right)^{2}/(m-2)}{\sum w_{j}\sum{w_{j}\hat{\gamma}}_{j}^{4}-\left( \sum w_{j}\hat{\gamma}_{j}^{2} \right)^{2}} \right)^{\frac{1}{2}}$$

(6)

$$se\left( \hat{\alpha}_{0} \right)=\left( \frac{\sum w_{j}\hat{\gamma}_{j}^{4}\sum w_{j}\left( \hat{\Gamma}_{j}\hat{\gamma}_{j}-\hat{\alpha}_{0}-\hat{\beta}\hat{\gamma}_{j}^{2} \right)^{2}/(m-2)}{\sum w_{j}\sum{w_{j}\hat{\gamma}}_{j}^{4}-\left( \sum w_{j}\hat{\gamma}_{j}^{2} \right)^{2}} \right)^{\frac{1}{2}}$$

(7)

#### 2. Weak instrument bias

To obtain a bias-corrected estimator, we start by writing the weighted least-squares estimator for $\beta$ in terms of the unobserved gene-exposure effects $\gamma_{j}$. This is the estimate that would be obtained if there were no weak instrument bias:

$$\hat{\beta}_{NOME}=\frac{\sum w_{j}\sum w_{j}\hat{\Gamma}_{j}\gamma_{j}^{3}-\sum w_{j}\hat{\Gamma}_{j}\gamma_{j}\sum w_{j}\gamma_{j}^{2}}{\sum w_{j}\sum{w_{j}\gamma}_{j}^{4}-\left( \sum w_{j}\gamma_{j}^{2} \right)^{2}}$$

(8)

We now approximate each sum in terms of observed quantities $\hat{\gamma}_{j}$ and $\sigma_{X_{j}}$. In the numerator,

$$w_{j}\hat{\Gamma}_{j}\gamma_{j}^{3}=w_{j}\hat{\Gamma}_{j}\left( \hat{\gamma}_{j}-\epsilon_{X_{j}} \right)^{3}=w_{j}\hat{\Gamma}_{j}\left( \hat{\gamma}_{j}^{3}-3\hat{\gamma}_{j}^{2}\epsilon_{X_{j}}+3\hat{\gamma}_{j}\epsilon_{X_{j}}^{2}-\epsilon_{X_{j}}^{3} \right)$$

$$=w_{j}\hat{\Gamma}_{j}\left( \hat{\gamma}_{j}^{3}-3\left( \gamma_{j}+\epsilon_{X_{j}} \right)^{2}\epsilon_{X_{j}}+3\left( \gamma_{j}+\epsilon_{X_{j}} \right)\epsilon_{X_{j}}^{2}-\epsilon_{X_{j}}^{3} \right)$$

$$=w_{j}\hat{\Gamma}_{j}\left( \hat{\gamma}_{j}^{3}-3\left( \gamma_{j}^{2}+2\gamma_{j}\epsilon_{X_{j}}+\epsilon_{X_{j}}^{2} \right)\epsilon_{X_{j}}+3\left( \gamma_{j}+\epsilon_{X_{j}} \right)\epsilon_{X_{j}}^{2}-\epsilon_{X_{j}}^{3} \right)$$

(9)

Since $E\left( \epsilon_{X_{j}} \right)=E\left( \epsilon_{X_{j}}^{3} \right)=0$ and $E\left( \epsilon_{X_{j}}^{2} \right)=\sigma_{Xj}^{2}$, all independently of $\Gamma_{j}\gamma_{j}$, we have

$$\sum w_{j}\hat{\Gamma}_{j}\gamma_{X_{j}}^{3}\approx\sum w_{j}\hat{\Gamma}_{j}\left( \hat{\gamma}_{j}^{3}-3\hat{\gamma}_{j}\sigma_{Xj}^{2} \right)$$

(10)

Similarly

$$w_{j}\hat{\Gamma}_{j}\gamma_{j}=w_{j}\hat{\Gamma}_{j}\left( \hat{\gamma}_{j}-\epsilon_{X_{j}} \right)$$

(11)

$$\sum w_{j}\hat{\Gamma}_{j}\gamma_{j}\approx\sum w_{j}\hat{\Gamma}_{j}\hat{\gamma}_{j}$$

(12)

and

$$w_{j}\gamma_{j}^{2}=w_{j}\left( \hat{\gamma}_{j}-\epsilon_{X_{j}} \right)^{2}=w_{j}\left( \hat{\gamma}_{j}^{2}-2\hat{\gamma}_{j}\epsilon_{X_{j}}+\epsilon_{X_{j}}^{2} \right)$$

$$=w_{j}\left( \hat{\gamma}_{j}^{2}-2\left( \gamma_{j}+\epsilon_{X_{j}} \right)\epsilon_{X_{j}}+\epsilon_{X_{j}}^{2} \right)$$

(13)

$$\sum w_{j}\gamma_{j}^{2}\approx\sum w_{j}\left( \hat{\gamma}_{j}^{2}-\sigma_{Xj}^{2} \right)$$

(14)

In the denominator,

$${w_{j}\gamma}_{j}^{4}=w_{j}\left( \hat{\gamma}_{j}-\epsilon_{X_{j}} \right)^{4}=w_{j}\left( \hat{\gamma}_{j}^{4}-4\hat{\gamma}_{j}^{3}\epsilon_{X_{j}}+6\hat{\gamma}_{j}^{2}\epsilon_{X_{j}}^{2}-4\hat{\gamma}_{j}\epsilon_{X_{j}}^{3}+\epsilon_{X_{j}}^{4} \right)$$

$$=w_{j}\left( \hat{\gamma}_{j}^{4}-4\left( \gamma_{j}+\epsilon_{X_{j}} \right)^{3}\epsilon_{X_{j}}+6(\left( \gamma_{j}+\epsilon_{X_{j}} \right)^{2}\epsilon_{X_{j}}^{2}-4\left( \gamma_{j}+\epsilon_{X_{j}} \right)\epsilon_{X_{j}}^{3}+\epsilon_{X_{j}}^{4} \right)$$

$$=w_{j}\left( \hat{\gamma}_{j}^{4}-4\left( \gamma_{j}^{3}+3\gamma_{j}^{2}\epsilon_{X_{j}}+3\gamma_{j}\epsilon_{X_{j}}^{2}+\epsilon_{X_{j}}^{3} \right)\epsilon_{X_{j}}+6\left( \gamma_{j}^{2}+2\gamma_{j}\epsilon_{X_{j}}+\epsilon_{X_{j}}^{2} \right)\epsilon_{X_{j}}^{2}-4\left( \gamma_{j}+\epsilon_{X_{j}} \right)\epsilon_{X_{j}}^{3}+\epsilon_{X_{j}}^{4} \right)$$

(15)

Since $E\left( \epsilon_{j}^{4} \right)=3\sigma_{Xj}^{2}$,

$$\sum{w_{j}\gamma}_{j}^{4}\approx\sum w_{j}\left( \hat{\gamma}_{j}^{4}-6\gamma_{j}^{2}{\sigma_{X}}_{j}^{2}-3\sigma_{Xj}^{4} \right)$$

$$=\sum w_{j}\left( \hat{\gamma}_{j}^{4}-6\left( \hat{\gamma}_{j}^{2}-2\hat{\gamma}_{j}\epsilon_{X_{j}}+\epsilon_{X_{j}}^{2} \right){\sigma_{X}}_{j}^{2}-3\sigma_{Xj}^{4} \right)$$

$$=\sum w_{j}\left( \hat{\gamma}_{j}^{4}-6\left( \hat{\gamma}_{j}^{2}-2\left( \gamma_{j}+\epsilon_{X_{j}} \right)\epsilon_{X_{j}}+\epsilon_{X_{j}}^{2} \right){\sigma_{X}}_{j}^{2}-3\sigma_{Xj}^{4} \right)$$

$$\approx\sum w_{j}\left( \hat{\gamma}_{j}^{4}-6\hat{\gamma}_{j}^{2}{\sigma_{X}}_{j}^{2}+3\sigma_{Xj}^{4} \right)$$

(16)

Although we have approximated each term in $\hat{\beta}_{NOME}$, this does not prove that the resulting estimator is consistent for $\beta$. We will study the accuracy of $\hat{\beta}_{NOME}$ in simulations.

To estimate a standard error, we need the variance of our approximation to $\hat{\beta}_{NOME}$, taken over an appropriate sampling distribution. We take the same approach as IVW and MR-Egger by conditioning on the observed predictors $\hat{\gamma}_{j}^{2}$ and sampling over the residuals $\alpha_{j}\hat{\gamma}_{j}+\epsilon_{Y_{j}}\hat{\gamma}_{j}$. As the denominator in $\hat{\beta}_{NOME}$ is fixed, conditional on $\hat{\gamma}_{j}^{2}$, we just need the variance of the numerator, which will be divided by the square of the denominator. Thus we need

$$var\left( \sum w_{j}\sum w_{j}\hat{\Gamma}_{j}\left( \hat{\gamma}_{j}^{3}-3\hat{\gamma}_{j}\sigma_{Xj}^{2} \right)-\sum w_{j}\hat{\Gamma}_{j}\hat{\gamma}_{j}\sum w_{j}\left( \hat{\gamma}_{j}^{2}-\sigma_{Xj}^{2} \right) \right)=var\left( \sum w_{j}\sum w_{j}\hat{\Gamma}_{j}\left( \hat{\gamma}_{j}^{3}-3\hat{\gamma}_{j}\sigma_{Xj}^{2} \right) \right)+var\left( \sum w_{j}\hat{\Gamma}_{j}\hat{\gamma}_{j}\sum w_{j}\left( \hat{\gamma}_{j}^{2}-\sigma_{Xj}^{2} \right) \right)-2cov\left( \sum w_{j}\sum w_{j}\hat{\Gamma}_{j}\left( \hat{\gamma}_{j}^{3}-3\hat{\gamma}_{j}\sigma_{Xj}^{2} \right),\sum w_{j}\hat{\Gamma}_{j}\hat{\gamma}_{j}\sum w_{j}\left( \hat{\gamma}_{j}^{2}-\sigma_{Xj}^{2} \right) \right)$$

$$=\left( \sum w_{j} \right)^{2}var\left( \sum w_{j}\hat{\Gamma}_{j}\left( \hat{\gamma}_{j}^{3}-3\hat{\gamma}_{j}\sigma_{Xj}^{2} \right) \right)+\left( \sum w_{j}\left( \hat{\gamma}_{j}^{2}-\sigma_{Xj}^{2} \right) \right)^{2}var\left( \sum w_{j}\hat{\Gamma}_{j}\hat{\gamma}_{j} \right)-2\sum w_{j}\sum w_{j}\left( \hat{\gamma}_{j}^{2}-\sigma_{Xj}^{2} \right)cov\left( \sum w_{j}\hat{\Gamma}_{j}\left( \hat{\gamma}_{j}^{3}-3\hat{\gamma}_{j}\sigma_{Xj}^{2} \right),\sum w_{j}\hat{\Gamma}_{j}\hat{\gamma}_{j} \right)$$

(17)

Assuming that SNPs are in linkage equilibrium, with independent effect sizes,

$$=\left( \sum w_{j} \right)^{2}\sum w_{j}^{2}\left( \hat{\gamma}_{j}^{2}-3\sigma_{Xj}^{2} \right)^{2}var(\hat{\Gamma}_{j}\hat{\gamma}_{j})+\left( \sum w_{j}\left( \hat{\gamma}_{j}^{2}-\sigma_{Xj}^{2} \right) \right)^{2}\sum w_{j}^{2}var\left( \hat{\Gamma}_{j}\hat{\gamma}_{j} \right)-2\sum w_{j}\sum w_{j}\left( \hat{\gamma}_{j}^{2}-\sigma_{Xj}^{2} \right)\sum w_{j}^{2}\left( \hat{\gamma}_{j}^{2}-3\sigma_{Xj}^{2} \right)var\left( \hat{\Gamma}_{j}\hat{\gamma}_{j} \right)$$

(18)

To estimate $var\left( \hat{\Gamma}_{j}\hat{\gamma}_{j} \right)$, note that

$$var\left( \hat{\Gamma}_{j}\hat{\gamma}_{j} \right)=E\left( \left( \hat{\Gamma}_{j}\hat{\gamma}_{j}-E\left( \hat{\Gamma}_{j}\hat{\gamma}_{j} \right) \right)^{2} \right)$$

$$=\hat{\gamma}_{j}^{2}E\left( \left( \hat{\Gamma}_{j}-\hat{\gamma}_{j}^{-1}E\left( \hat{\Gamma}_{j}\hat{\gamma}_{j} \right) \right)^{2} \right)$$

(19)

We estimate $E\left( \hat{\Gamma}_{j}\hat{\gamma}_{j} \right)$ by the fitted value of the standard GRIP regression (without weak instrument adjustment). For the outer expectation we take an empirical estimate by averaging across SNPs $j$, assuming VICE.

#### 3. Further data results

The multivariate analyses for LDL and HDL cholesterol on coronary heart disease are shown in Tables A and B in S1 Text respectively.

|  | Coding | $\exp\left( \hat{\beta} \right)$ (95% CI)_ | $se(\hat{\beta})$ | $P$ | Intercept $P$ |
| --- | --- | --- | --- | --- | --- |
| Univariate |  |  |  |  |  |
| IVW |  | 1.53 (1.41 – 1.66) | 0.0410 | 2.1e-25 |  |
| MR-Egger |  | 1.57 (1.42 – 1.74) | 0.0528 | 5.2e-15 | 0.471 |
| MR-GRIP |  | 1.54 (1.42 – 1.66) | 0.0410 | 2.3e-20 | 0.193 |
| Multivariate |  |  |  |  |  |
| IVW |  | 1.47 (1.36 – 1.60) | 0.0412 | 2.3e-17 |  |
| MR-Egger | LDL | 1.55 (1.40 – 1.71) | 0.0509 | 5.1e-15 | 0.111 |
|  | HDL | 1.48 (1.37 – 1.60) | 0.0403 | 4.0e-18 | 0.00342 |
|  | Triglycerides | 1.44 (1.33 – 1.56) | 0.0418 | 1.7e-15 | 0.020 |
| MR-GRIP | LDL | 1.48 (1.36 – 1.60) | 0.0411 | 1.6e-17 | 0.170 |
|  | HDL | 1.47 (1.36 – 1.60) | 0.0412 | 2.9e-17 | 0.970 |
|  | Triglycerides | 1.48 (1.36 – 1.60) | 0.0412 | 2.0e-17 | 0.298 |

**Table A.** Estimated causal effects of LDL cholesterol on coronary heart disease. $\exp(\hat{\beta})$, estimated odds ratio per 1-sd increase in inverse-normal transformed LDL. $se(\hat{\beta})$, standard error of log odds ratio. Coding, for MR-Egger, all-positive coding with respect to listed trait; for MR-GRIP, equation (9) multiplied by SNP-exposure effects for listed trait.

|  | Coding | $\exp\left( \hat{\beta} \right)$ (95% CI)_ | $se(\hat{\beta})$ | $P$ | Intercept $P$ |
| --- | --- | --- | --- | --- | --- |
| Univariate |  |  |  |  |  |
| IVW |  | 0.874 (0.793 – 0.962) | 0.0492 | 0.0067 |  |
| MR-Egger |  | 0.985 (0.865 – 1.11) | 0.0615 | 0.810 | 0.00200 |
| MR-GRIP |  | 0.873 (0.793 – 0.962) | 0.0494 | 0.0069 | 0.979 |
| Multivariate |  |  |  |  |  |
| IVW |  | 0.955 (0.880 – 1.04) | 0.0417 | 0.272 |  |
| MR-Egger | LDL | 0.960 (0.885 – 1.04) | 0.0416 | 0.326 | 0.111 |
|  | HDL | 1.03 (0.94 – 1.14) | 0.0490 | 0.487 | 0.00342 |
|  | Triglycerides | 0.974 (0.897 – 1.06) | 0.0420 | 0.528 | 0.020 |
| MR-GRIP | LDL | 0.958 (0.883 – 1.04) | 0.0416 | 0.302 | 0.170 |
|  | HDL | 0.955 (0.880 – 1.04) | 0.0418 | 0.274 | 0.970 |
|  | Triglycerides | 0.959 (0.884 – 1.04) | 0.0417 | 0.324 | 0.298 |

**Table B.** Estimated causal effects of HDL cholesterol on coronary heart disease. $\exp(\hat{\beta})$, estimated odds ratio per 1-sd increase in inverse-normal transformed HDL. $se(\hat{\beta})$, standard error of log odds ratio. Coding, for MR-Egger, all-positive coding with respect to listed trait; for MR-GRIP, equation (9) multiplied by SNP-exposure effects for listed trait.

#### 4. Further simulation results

We repeated the simulations, taking SNP-exposure effects  $\hat{\gamma}_{j}$ from the BMI data example. Results are qualitatively similar to the urate example, with MR-GRIP unbiased under scenario 1 (balanced pleiotropy) and scenario 4 (VICE) and showing a bias that is intermediate between IVW and MR-Egger in scenario 2 (all-positive InSIDE) and scenario 3 (InSIDE under original coding).

| Scenario | Proportion invalid | IVW | MR-Egger | MR-GRIP | MR-GRIP weak | Weighted median | Mode-based estimator |
| --- | --- | --- | --- | --- | --- | --- | --- |
| 1 | 0.3 | 0.197 | 0.182 | 0.186 | 0.202 | 0.192 | 0.194 |
|  | 1 | 0.197 | 0.180 | 0.186 | 0.202 | 0.191 | 0.193 |
| 2 | 0.3 | 0.389 | 0.179 | 0.279 | 0.269 | 0.303 | 0.266 |
|  | 1 | 0.854 | 0.180 | 0.508 | 0.435 | 0.724 | 0.604 |
| 3 | 0.3 | 0.213 | 0.260 | 0.234 | 0.261 | 0.214 | 0.229 |
|  | 1 | 0.254 | 0.454 | 0.353 | 0.411 | 0.387 | 0.496 |
| 4 | 0.3 | 0.225 | 0.161 | 0.189 | 0.201 | 0.208 | 0.203 |
|  | 1 | 0.293 | 0.114 | 0.198 | 0.200 | 0.251 | 0.229 |

**Table C.** Mean estimates of $\beta=0.2$ with 96 SNPs as instruments. SNP-exposure effects as for the BMI data example (Table B in S1 Data). Scenarios described in main text. Proportion invalid, proportion of SNPs with direct pleiotropic effects on outcome. MR-GRIP weak, MR-GRIP with adjustment for weak instruments.

| Scenario | Proportion invalid | Standard error | IVW | MR-Egger | MR-GRIP | MR-GRIP weak | Weighted median | Mode-based estimator |
| --- | --- | --- | --- | --- | --- | --- | --- | --- |
| 1 | 0.3 | Analytic | 0.0407 | 0.0998 | 0.0614 | 0.128 |  |  |
|  |  | Empirical | 0.0402 | 0.0961 | 0.0608 | 0.0865 | 0.0578 | 0.0800 |
|  | 1 | Analytic | 0.0414 | 0.101 | 0.0629 | 0.130 |  |  |
|  |  | Empirical | 0.0407 | 0.100 | 0.0627 | 0.0899 | 0.0597 | 0.0830 |
| 2 | 0.3 | Analytic | 0.0528 | 0.127 | 0.0809 | 0.166 |  |  |
|  |  | Empirical | 0.0428 | 0.129 | 0.0792 | 0.117 | 0.0737 | 0.101 |
|  | 1 | Analytic | 0.0515 | 0.101 | 0.0638 | 0.195 |  |  |
|  |  | Empirical | 0.0428 | 0.0995 | 0.0647 | 0.0897 | 0.0620 | 0.106 |
| 3 | 0.3 | Analytic | 0.0562 | 0.138 | 0.0867 | 0.172 |  |  |
|  |  | Empirical | 0.0528 | 0.130 | 0.0817 | 0.119 | 0.0728 | 0.102 |
|  | 1 | Analytic | 0.0842 | 0.206 | 0.130 | 0.248 |  |  |
|  |  | Empirical | 0.0422 | 0.117 | 0.0783 | 0.0938 | 0.0873 | 0.120 |
| 4 | 0.3 | Analytic | 0.0409 | 0.100 | 0.0617 | 0.128 |  |  |
|  |  | Empirical | 0.0393 | 0.0970 | 0.0611 | 0.0871 | 0.0571 | 0.0791 |
|  | 1 | Analytic | 0.0415 | 0.100 | 0.0619 | 0.133 |  |  |
|  |  | Empirical | 0.0400 | 0.0985 | 0.0621 | 0.0877 | 0.0575 | 0.0797 |

**Table D.** Mean analytic standard errors, and empirical standard deviations of point estimates in simulations of Table C in S1 Text.

| Scenario | Proportion invalid | MR-Egger | MR-GRIP | Correlation |
| --- | --- | --- | --- | --- |
| 1 | 0.3 | 0.0515 | 0.0599 | 0.880 |
|  | 1 | 0.0556 | 0.0659 | 0.880 |
| 2 | 0.3 | 0.470 | 0.466 | 0.955 |
|  | 1 | 1.00 | 1.00 | 0.903 |
| 3 | 0.3 | 0.0498 | 0.0593 | 0.863 |
|  | 1 | 0.0014 | 0.0086 | 0.832 |
| 4 | 0.3 | 0.734 | 0.726 | 0.948 |
|  | 1 | 1.00 | 1.00 | 0.885 |

**Table E.** Power (at $P\leq0.05)$ of the intercept tests of MR-Egger and MR-GRIP in simulations of Table C in S1 Text. $P$-values obtained from the ratio of point estimate to analytic standard error, assuming a standard normal distribution. Correlation, Spearman correlation between $P$-values of the two tests.

For the weak instrument simulation (Table 5 in the main text), the adjusted MR-GRIP showed greater numerical instability than the urate example (Fig S1 and Fig S2).
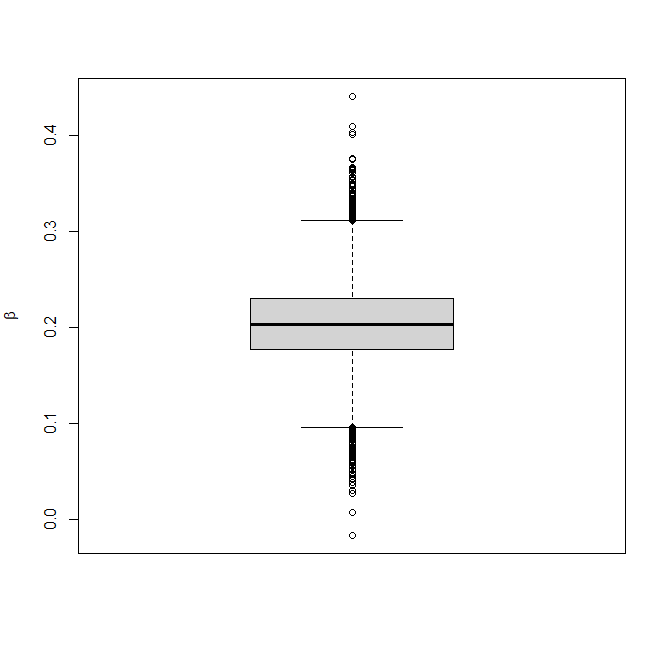

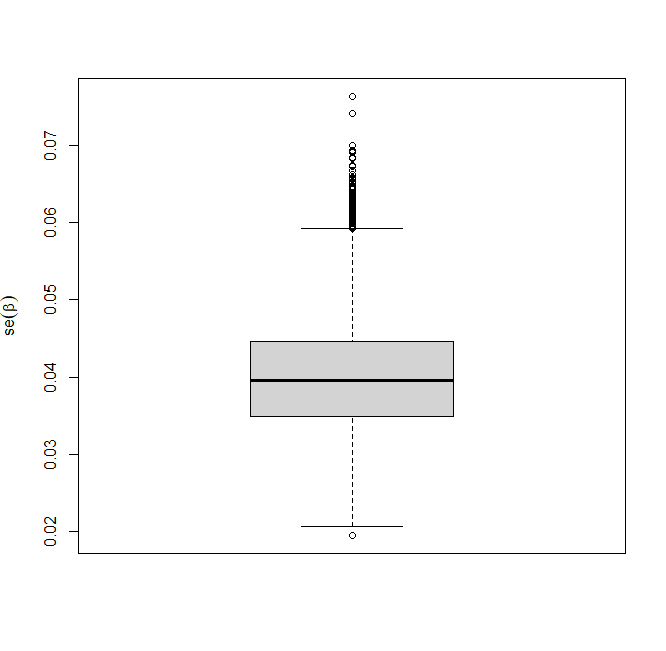
**Fig A.** Boxplots of causal effect estimates (left) and standard errors (right) for the weak instrument adjusted MR-GRIP in the urate simulation.

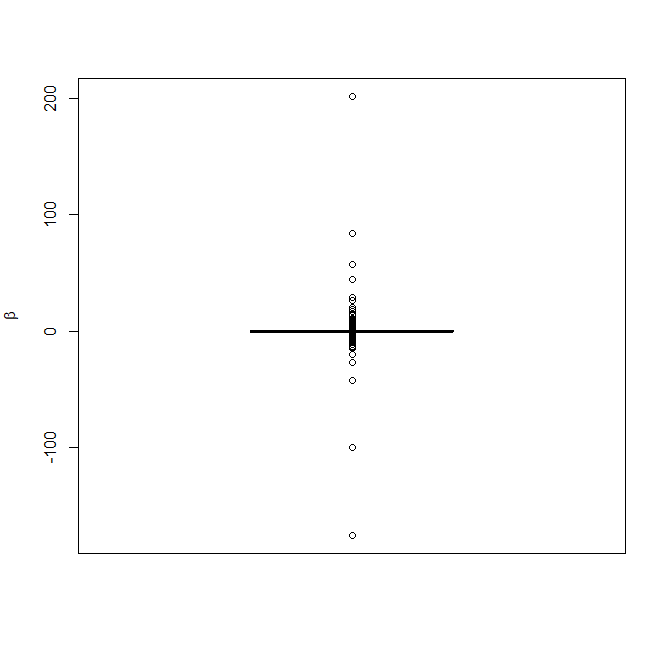

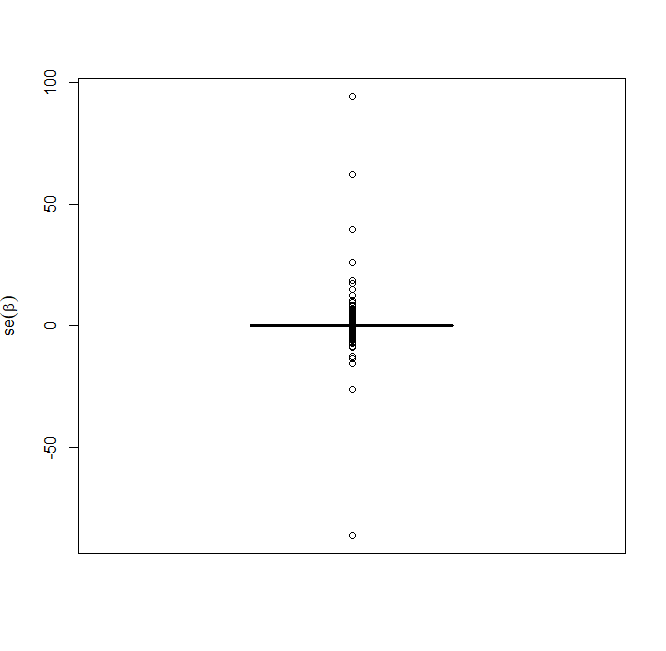

**Fig B.** Boxplots of causal effect estimates (left) and standard errors (right) for the weak instrument adjusted MR-GRIP in the BMI simulation.
